# Reduced Reflex Autonomic Responses following Intradetrusor OnabotulinumtoxinA Injections: A pre/post study in Individuals with Cervical and Upper Thoracic Spinal Cord Injury

**DOI:** 10.1101/2021.04.24.21256011

**Authors:** Tristan W. Dorey, Matthias Walter, Andrei V Krassioukov

## Abstract

**Importance:** Urodynamic studies (UDS) can provoke autonomic dysreflexia (AD) in individuals with spinal cord injury (SCI) at and above the sixth thoracic (T6) spinal segment potentially leading to profound vagally mediated heart rate reductions. Intradetrusor onabotulinumtoxinA injections have been shown to reduce AD events during UDS in this cohort but evidence is lacking whether this treatment can improve reflex autonomic responses.

**Objective:** To assess the effects of intradetrusor onabotulinumtoxinA injections on heart rate variability (HRV) responses to UDS in individuals, 18-65 years of age, with chronic (>1-year) SCI at or above T6 with confirmed neurogenic detrusor overactivity and AD during UDS.

**Design, Setting, and Participants:** This cohort study used participants from our recent prospective phase IV clinical trial. Individuals were screened at an academic medical centre between November 2014 and December 2019. After enrollment, participants underwent UDS prior to (i.e., baseline) and one month after intradetrusor onabotulinumtoxinA injections (post-treatment).

**Interventions:** OnabotulinumtoxinA (200 U) was injected into the detrusor muscle at 20 sites (10 U/site).

**Main Outcomes and Measures:** Continuous electrocardiogram (ECG) and blood pressure (BP) recordings were used to assess RR-interval, time and frequency domain metrics of HRV (a surrogate marker of autonomic nervous system activity), and AD pre- and post-intervention.

**Results:** A total of 19 participants with SCI (5 women; mean [SD] age 42 [8.3] years) with complete ECG and BP data sets were suitable for autonomic analysis. During baseline UDS, an increase in RR-interval (adjusted mean difference, -0.08; 95% CI, -0.14 to -0.03; *P*=0.002) as well as time and frequency domain metrics of HRV were detected. Vagally mediated increases in high frequency (HF) power during UDS were larger in participants with cervical SCI compared to upper thoracic SCI (adjusted mean difference, 20.3; 95% CI, 3.3 to 37.2; *P*=0.013). Intradetrusor onabotulinumtoxinA injections significantly reduced time domain metrics of HRV and HF power (adjusted mean difference, 9.1; 95% CI, 3.1 to 15.1, *P*<0.01) responses to UDS across all participants.

**Conclusions and Relevance:** Changes in HRV during UDS could be a potential indicator of improved autonomic cardiovascular function following interventions such as intradetrusor onabotulinumtoxinA injections.

**Trial Registration:** ClinicalTrials.gov Identifier: NCT02298660

## INTRODUCTION

Spinal cord injury (SCI) results in damage to descending autonomic pathways causing a wide array of autonomic dysfunctions.^1,2^ Autonomic dysreflexia (AD) is a potentially life-threatening condition characterized by an abrupt increase in systolic blood pressure (SBP) ≥20 mmHg due to innocuous or noxious stimuli below the level of injury.^3,4^ Neurogenic detrusor overactivity (NDO) is the leading cause of AD events in individuals with SCI and requires routine urodynamic studies (UDS) for surveillance and management of lower urinary tract (LUT) function.^5,6^ AD episodes can be made more serious due to reflexive vagal activation that causes profound bradycardia leading to dysrhythmias, such as atrial fibrillation, sinus pauses, or AV node block.^7–9^

Considering the aforementioned, and the potential risk of cardiovascular complications associated with AD during UDS in individuals with SCI^10^, the need for safe and effective treatments that address both NDO and subsequent autonomic consequences is paramount. Recently, our group published data from a phase IV clinical trial demonstrating that intradetrusor onabotulinumtoxinA injections are effective at ameliorating AD during UDS while improving LUT function and overall quality of life in individuals with cervical and upper thoracic SCI.^11,12^ Despite this finding, it is still unknown if intradetrusor onabotulinumtoxinA injections can also improve reflex vagal responses to bladder filling during UDS in individuals with SCI.

Heart rate variability (HRV) is a powerful tool used to non-invasively assess autonomic regulation of the cardiovascular system. Reductions in the beat-to-beat variation of R-R interval on the electrocardiogram (ECG) are associated with worsened overall health status in a number of disease conditions due to increasing sympathetic tone.^13–15^ In SCI, HRV provides valuable and reliable feedback on the integrity and responsiveness of autonomic pathways in response to external stimuli that is graded by neurological level of injury (NLI).^16–18^ Specifically, frequency domain analysis of HRV has been shown to be a powerful tool for predicting clinical cardiovascular dysfunction in individuals with SCI.^16^ In the present study, we use HRV to assess autonomic nervous system responses to bladder filling during UDS in individuals with cervical and upper thoracic SCI that underwent the trial. We hypothesized that intradetrusor onabotulinumtoxinA injections would improve reflex autonomic regulation of the cardiovascular system in SCI patient undergoing UDS.

## METHODS

### Study design

This prospective phase IV clinical trial using a pre-post study design was approved by the University of British Columbia Clinical Research Ethics Board and registered at clinicaltrials.gov (identifier NCT02298660).

### Participants

Between November 2014 and December 2019, 55 individuals with chronic SCI (>1-year post-injury) at or above T6 were screened based on inclusion and exclusion criteria reported previously.^11^ 34 individuals with chronic SCI at T6 or above with confirmed history of AD and NDO were included and assigned to undergo intradetrusor onabotulinumtoxinA injections (200 IU) intended to improve LUT function and ameliorate bladder-related AD. One month following intradetrusor onabotulinumtoxinA injections, UDS were repeated. Of these participants, 13 individuals had incomplete ECG recordings (i.e. not suitable enough for analysis) and two participants were excluded from analysis due to the presence of arrhythmic events that prevented HRV analysis (**Figure 1**).

**Figure 1:**
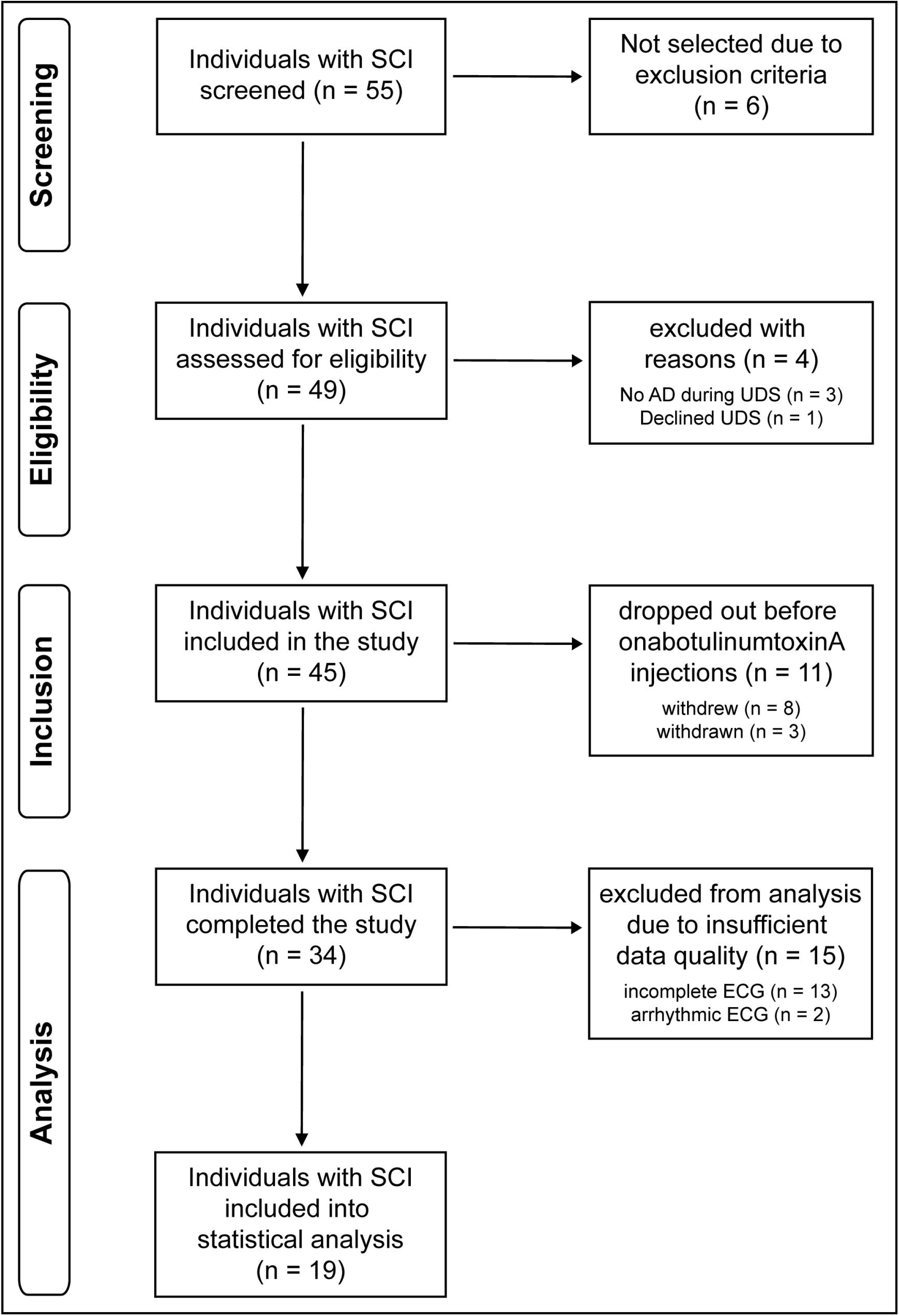
Trial flow diagram. AD; autonomic dysreflexia, ECG; electrocardiogram, SCI; spinal cord injury, UDS; urodynamic study.

### Study Assessments

NLI and completeness (i.e. AIS grade) of SCI were classified according to the International Standards for Neurological Classification of SCI (ISNCSCI).^19^ All Urodynamics (UDS) were performed with a Aquarius TT (Laborie Model 94-R03-BT, Montreal, Quebec, Canada) in accordance with the International Continence Society.^20^ Concurrent to UDS, we continuously recorded beat-by-beat blood pressure, via finger photoplethysmography (Finometer PRO, Finapres Medical Systems, Amsterdam, Netherlands) corrected to brachial pressure (CARESCAPE V100, GE Healthcare, Milwaukee, WI, USA), and one-lead electrocardiogram (eML 132; ADInstruments, Colorado Springs, CO, USA) for heart rate (HR) in order to detect autonomic dysreflexia^21,22^ and to analyze HRV (see below).

### Heart Rate Variability Analysis

HRV was assessed using time and frequency domain analysis in accordance with the European task force HRV guidelines^23^ and as described elsewhere.^24,25^ Briefly, ECG filtering and R-wave detection was performed using LabChart (ADInstruments, V8, Colorado Springs, CO, USA). The RR interval time series were obtained over 4-5-minute segments during supine rest and UDS at baseline and post-treatment. Due to the need for at least 4 minutes of stationary recording for accurate frequency domain analysis^22^, UDS measurements were taken from the 4-5 minutes immediately before strong desire to void. Each segment was manually examined to ensure stationary and stable sinus rhythm with no trend (i.e. average increase or decrease) in RR interval over the segment. If participants did not have stable sinus rhythm at baseline or during UDS, as classified according to American Heart Association (AHA) guidelines^26^, they were excluded from analysis (**Figure 1**).

Time domain parameters were calculated for each segment which included the standard deviation of all normal RR intervals (SDNN) and the root mean squared of successive differences in RR interval (RMSSD). SDNN and RMSSD represent general variability and parasympathetically driven changes in RR interval respectively.^27^ The same segments were subsequently used for frequency domain analysis using Welch’s method with 50% windowing.^24,25^ The periodogram of each segment was then calculated using the Fourier transformation. Total power of each periodogram was measured as a total index of HRV, which is determined by the integral over the entire frequency range. The low frequency (LF) and high frequency (HF) components were then extracted. The LF oscillations in HR (0.1-1.5 Hz) are regulated by both the sympathetic and parasympathetic nervous systems while the HF component (1.5-5 Hz) is predominantly mediated by the phasic activity of the parasympathetic nervous system.^27^ LF and HF powers were normalized by dividing the power by the total power minus very low frequency (VLF) power and multiplied by 100.

Beating interval variability was also assessed using non-linear Poincaré plot analysis. The standard deviations (SD1 and SD2) of each plot were calculated from the RR-interval time series using the following equations: SD1^2^=2[SD(RRn-RRn+1]^2^ and SD2^2^=[SD(RR)]^2^-1/2[SD(RRn-RRn+1]^2^.

### Statistical Analysis

All data are presented as means with standard deviation [SD]. Statistical analysis was conducted using Prism version 9.0.0 (GraphPad Software, San Diego). All data were tested for normality and equal variance by a Kolmogorov-Smirnov test and an F-test respectively. Data were analyzed using two-way repeated measures ANOVA with Tukey post-hoc test as indicated in each figure legend and adjusted mean differences with 95% confidence intervals (CI) were reported in text for each comparison. The assumption of sphericity was tested using Mauchly’s test and the Greenhouse-Geisser correction factor to the degrees of freedom was used for all positive tests. Correlations between the change in HRV in response to bladder filling during UDS, and SBP changes during UDS and AD were performed using Pearson correlations. *P*<0.05 was considered to be significant.

## RESULTS

In total, 19 individuals (5 women; mean [SD]; age, 42 [8.3] years; time post-injury, 25.5 [13.5]) were included in the overall analysis (see participant demographics in **Table 1**). To perform injury-level-dependent analyses, all participants in the present cohort with upper thoracic SCI (n=6 mean [SD]; age, 44 [8.83] years; time-post-injury, 20 [13.7] years) were assigned age matched (within 5 years) cervical SCI controls for comparison (n=6 mean [SD] age, 44.3 [10.2] years; time-post-injury, 14.3 [13.7] years). Accordingly, no significant difference in age was observed between the two groups (mean difference, -0.33; 95% CI-12.6 to 12.0; *P*=0.95). In the whole cohort, majority of participants had motor-complete SCI in accordance with AIS (A=8, B=8, C=3).

**Table 1:** Patient Characteristics

| ID | NLI | AIS | Age range* (yr) | Sex | TPI range* (yr) | History of OnabotulinumtoxinA injections |
| --- | --- | --- | --- | --- | --- | --- |
| 1 | C1 | C | 41-45 | M | 26-30 | Naive |
| 2 | C5 | B | 31-35 | M | 1-5 | Naive |
| 3 | C5 | C | 61-65 | M | 1-5 | Naive |
| 4 | C5 | B | 31-35 | M | 1-5 | Yes |
| 5 | C6 | A | 36-40 | F | 11-15 | Naive |
| 6 | C6 | B | 31-35 | F | 11-15 | Naive |
| 7 | C6 | C | 46-50 | F | 26-30 | Naive |
| 8 | C7 | A | 41-45 | F | 16-20 | Naive |
| 9 | C7 | B | 36-40 | M | 16-20 | Naive |
| 10 | C7 | B | 36-40 | M | 1-5 | Naive |
| 11 | C7 | B | 46-50 | M | 6-10 | Naive |
| 12 | C8 | A | 41-45 | M | 21-25 | Naive |
| 13 | C8 | B | 41-45 | M | 36-40 | Naive |
| 14 | T2 | A | 36-40 | M | 6-10 | Yes |
| 15 | T2 | A | 36-40 | M | 6-10 | Yes |
| 16 | T3 | B | 36-40 | M | 16-20 | Yes |
| 17 | T4 | A | 36-40 | F | 21-25 | Naive |
| 18 | T4 | A | 56-60 | M | 1-5 | Naive |
| 19 | T5 | A | 44-50 | M | 1-5 | Naive |
| **Mean** | C=13 | A=8 | 42.5 | M=14 | 25.5 | Yes=4 |
| **± SD** | T=6 | B=8<br>C=3 | ±8.3 | F=5 | ±13.5 | naïve=15 |

\* For information, such as age and time post injury, that would allow the study participant or their family, friends or neighbors to identify them, we chose to provide a 5-year range for age and time post injury, rather than revealing the actual numbers.
AIS; American spinal injury impairment scale, C; cervical, F: female, M: male, NLI; neurological level of injury, SD; standard deviation, T; thoracic, TPI; time post-injury, yr: years

First, we assessed time and frequency domain HRV responses to bladder filling during UDS. Overall, the majority of individuals (57%) presented with clinically defined bradycardia during UDS.^25^ RR-interval significantly increased in the whole cohort (adjusted mean difference, -0.09; 95% CI,-0.14 to -0.03 ;*P*=0.002 ;**Figure 2A and B**) and in individuals with cervical SCI (adjusted mean difference, -0.12; 95% CI, -0.23 to 0.02; *P*=0.01) in response to bladder filling during UDS. There was no change in RR-interval in response to bladder filling during UDS in individuals with thoracic SCI (adjusted mean difference, -0.06; 95% CI, -0.17 to 0.04, *P*=0.31). Bladder filling during UDS increased general variability as assessed by SDNN in the whole cohort (adjusted mean difference, -18.3; 95% CI, -29.2 to -7.5, *P*<0.001; **Figure 2C-D**) and in cervical SCI (adjusted mean difference, -27.6; 95% CI, -46.8 to -8.3, *P*<0.01) but not thoracic SCI (adjusted mean difference, -15.9; 95% CI, -35.2 to 3.4, *P*=0.13). Similarly, parasympathetically driven RMSSD was increased during UDS in the whole cohort (adjusted mean difference, -15.5; 95% CI, -25.7 to -5.2, *P*<0.01; **Figure 2E**) and in cervical SCI (adjusted mean difference, -19.9; 95% CI, -38.2 to -1.5, *P*<0.05). RMSSD was not significantly different during UDS in thoracic SCI (adjusted mean difference, -9.6; 95% CI, -27.9 to 8.7, *P*=0.47). Non-linear analysis of HRV assessed by Poincaré plot analysis (**Figure 2F**) demonstrated significant increases in SD1 (adjusted mean difference, -10.6; 95% CI, -17.9 to -3.3, *P*<0.01; **Figure 2G**) and SD2 (adjusted mean difference, -22.7; 95% CI, -38.7 to -6.8, *P*<0.05; **Figure 2H**) in the whole cohort. Cervical SCI also had significant increases in SD1 (adjusted mean difference, -14.2; 95% CI, -27.3 to -1.2, *P*<0.05) and SD2 (adjusted mean difference, -34.6; 95% CI, -63 to -6.3, *P*<0.05). SD1 did not differ in thoracic SCI (adjusted mean difference, -6.8; 95% CI, -19.9 to 6.2, *P*=0.48). While SD2 also did not differ in thoracic SCI (adjusted mean difference, -25.3; 95% CI, -53.7 to 3.1, *P*=0.09) during bladder filling, there was a large effect size (Cohen’s *d* = 0.91) following bladder filling. Spectral analysis of LF power decreased overall (adjusted mean difference, 8.3; 95% CI, 0.9 to 15.7, *P*<0.05; **Figure 2I-J**), in cervical SCI (adjusted mean difference, 13.9; 95% CI, 0.8 to 27.1, *P*<0.05), but did not change in thoracic SCI (adjusted mean difference, -4.2; 95% CI, -17.3 to 8.9, *P*=0.8) in response to bladder filling during UDS. Conversely, HF power increased overall (adjusted mean difference, -8.9; 95% CI, -16.2 to -1.5, *P*<0.05), in cervical SCI (adjusted mean difference, -13.9; 95% CI, -27 to -0.9, *P*<0.05), but did not change in thoracic SCI (adjusted mean difference, 4.2; 95% CI, -8.8 to 17.3, *P*=0.8).

**Figure 2:**
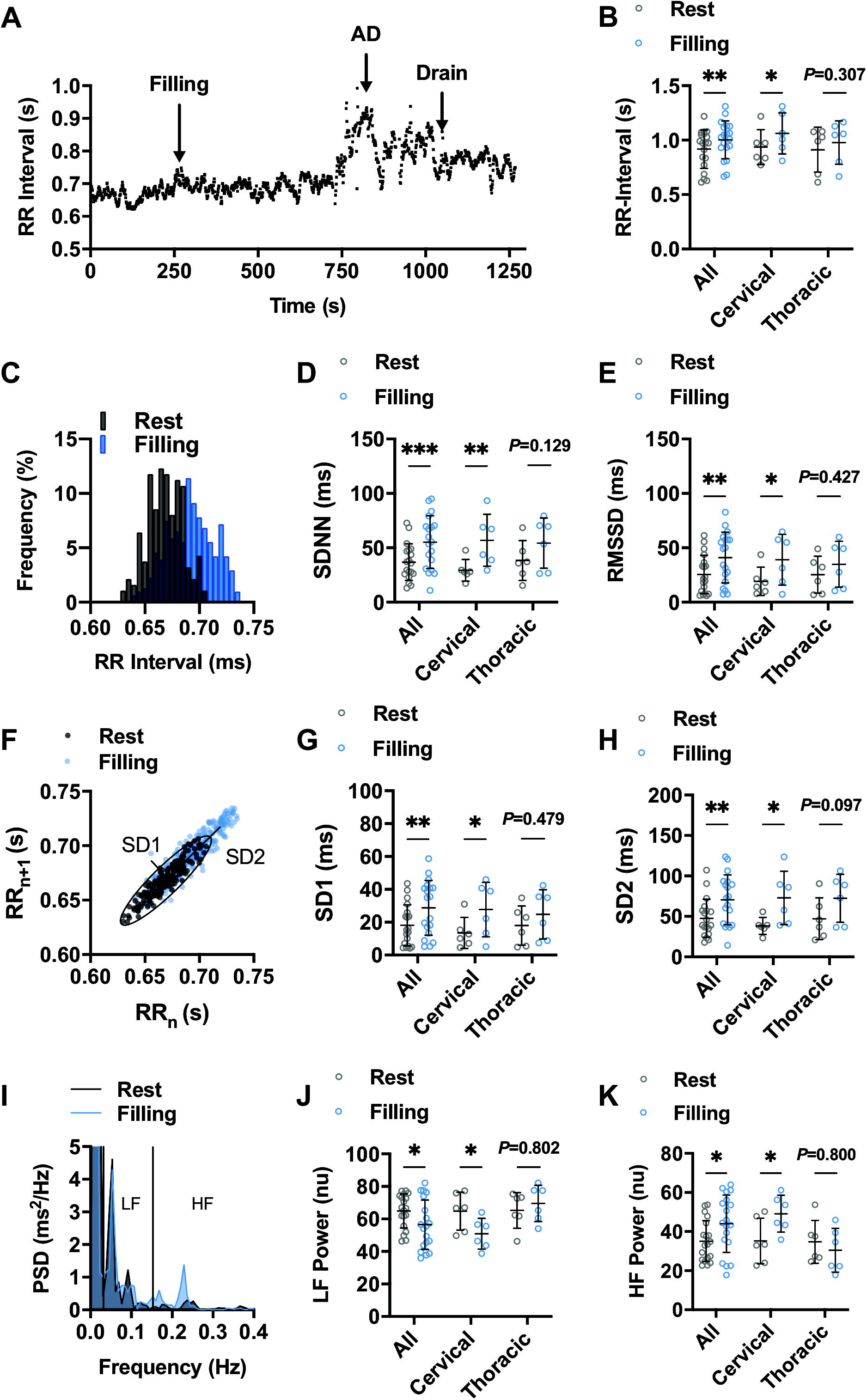
The effect of urodynamic study (UDS) on RR-interval and HR variability (HRV) in individuals with cervical (n=13) and upper thoracic (n=6) spinal cord injury (SCI). **A**. Representative RR-interval time series from participant 2 demonstrating prolongation of RR-interval with bladder filling and subsequent bradycardia during autonomic dysreflexia (AD) event. **B**. Summary RR-interval data at rest and following bladder filling in the whole cohort (all) and in injury-level-dependent subgroups. **C**. Representative RR histogram at rest and during bladder filling. **D and E**. Summary standard deviation of N-N intervals (SDNN; **D**) and root mean squared of successive differences (RMSSD; **E**) at rest and during bladder filling. **F-H**. Representative non-linear Poincaré plot analysis (**F**) and summary standard deviation 1 (SD1; **G**) and SD2 (**H**) at rest and during bladder filling. I-K. Representative power spectral density plots (I) and summary data for normalized low-frequency (LF; **J**) and high frequency (HF; **K**) power. \**P*<0.05, \*\**P<*0.01, \*\*\**P*<0.001 by 2-way ANOVA with Tukey post-hoc test.

One-month post-intradetrusor onabotulinumtoxinA injections, no differences were observed in the RR-interval response to bladder filling during UDS (adjusted mean difference, -0.03; 95% CI, -0.07 to 0.01, *P*=0.15; **Figure 3A**). There was no significant difference in RR-interval post-intradetrusor onabotulinumtoxinA injection compared to baseline UDS values (adjusted mean difference, 0.04; 95% CI, -0.002 to 0.08, *P*=0.07). Significant reductions in response to bladder filling were also observed in SDNN (adjusted mean difference, 13.4; 95% CI, 4.1 to 22.6, *P*<0.01; **Figure 3B**) and RMSSD (adjusted mean difference; 13.1; 95% CI, 3.5 to 22.6, *P*<0.01; **Figure 3C**) compared to baseline values. Intradetrusor onabotulinumtoxinA also significantly reduced nonlinear SD1 (adjusted mean difference, 9.1; 95% CI, 2,5 to 15.7, *P*<0.01; **Figure 3D**) and SD2 (adjusted mean difference, 15.9; 95% CI, 1.6 to 30.3, *P*<0.05; **Figure 3E**) compared to baseline UDS. Furthermore, post-treatment LF power during UDS was significantly increased (adjusted mean difference, -9.1; 95% CI, -15.1 to -3.1, *P*<0.01) while HF power was decreased (adjusted mean difference, 9.1; 95% CI, 3.1 to 15.1, *P*<0.01) compared to baseline UDS.

**Figure 3:**
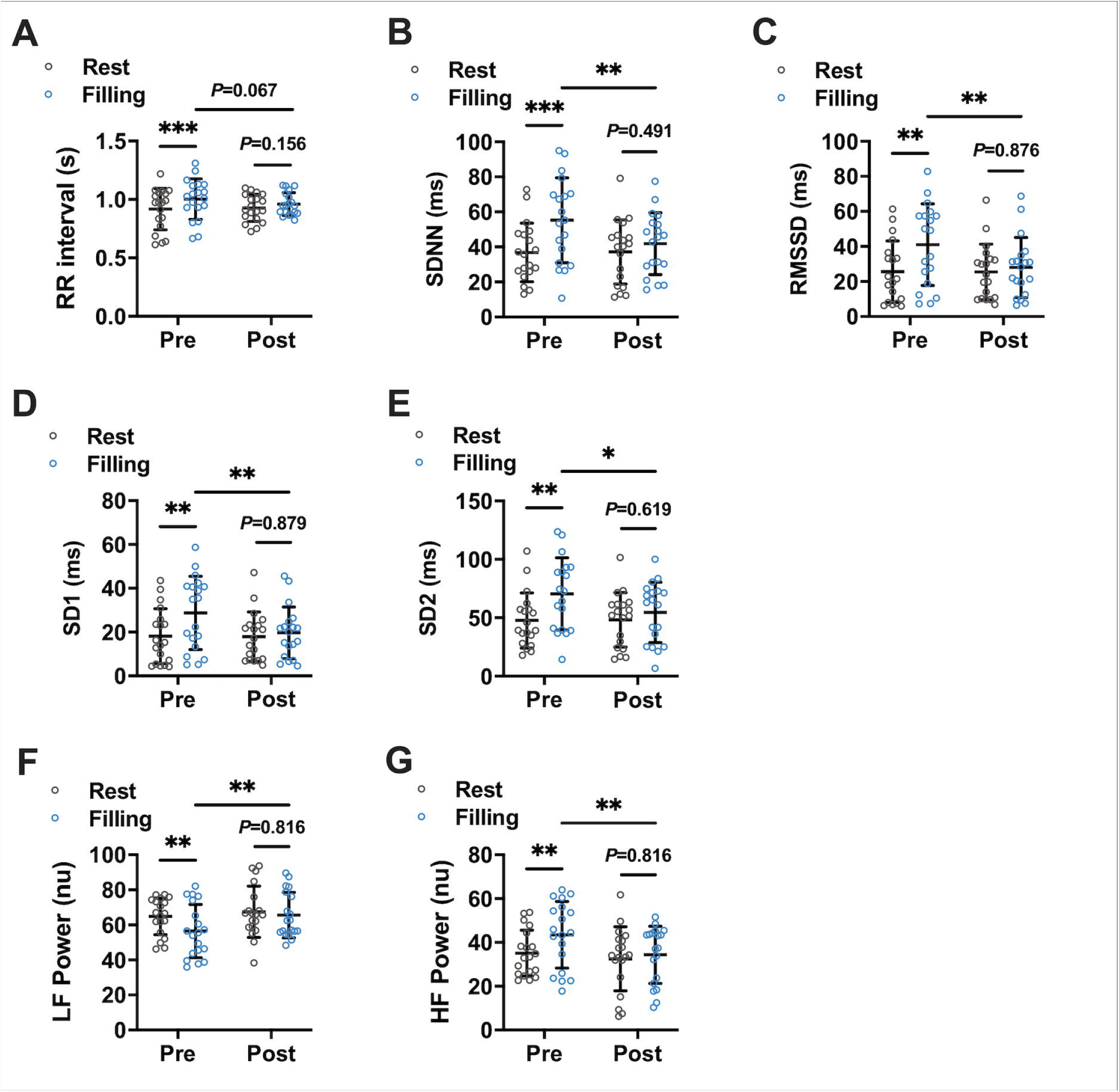
Effects of onabotulinumtoxinA treatment on HRV responses to UDS. Pre/post onabotulinumtoxinA treatment comparison of (**A**) RR-interval, (**B**) SDNN, (**C**) RMSSD, (**D**) SD1, (**E**) SD2, (**F**) normalized LF Power, and (**G**) normalized HF power responses to UDS at rest and during bladder filling. \**P*<0.05, \*\**P<*0.01, \*\*\**P*<0.001 by 2-way ANOVA with Tukey post-hoc test.

To assess if the changes in HRV were associated with SBP through the cardiovagal baroreflex, we correlated the change in HRV metrics during UDS for all participants baseline / post-treatment with the changes in SBP during UDS (ΔSBP_UDS_) at the same timepoint (**Table 2**). Significant correlations were present for ΔRR-interval (r=0.44 *P=*0.005), ΔRMSSD (r=0.47 *P=*0.003), ΔSD1 (r=0.47 *P=*0.003), ΔLF power (r=-0.59 *P<*0.0001), and ΔHF power (r=0.59 *P*<0.0001). We also examined the association between these metrics and the average change in SBP during AD (ΔSBP_AD_). Significant correlations were present for ΔRR-interval (r=0.35 *P=*0.03), ΔSDNN (r=0.33 *P=*0.04), ΔRMSSD (r=0.37 *P=*0.02), ΔSD1 (r=0.37 *P=*0.02), ΔLF power (r=-0.43 *P=*0.007), and ΔHF power (r=0.43 *P=*0.007). Furthermore, ΔRMSSD (r=0.33 *P=*0.04), ΔSD1 (r=0.33 *P=*0.04), ΔLF power (r=-0.43 *P=*0.006) and ΔHF power (r=0.43 *P=*0.006) were significantly associated with maximal SBP during AD.

**Table 2:**
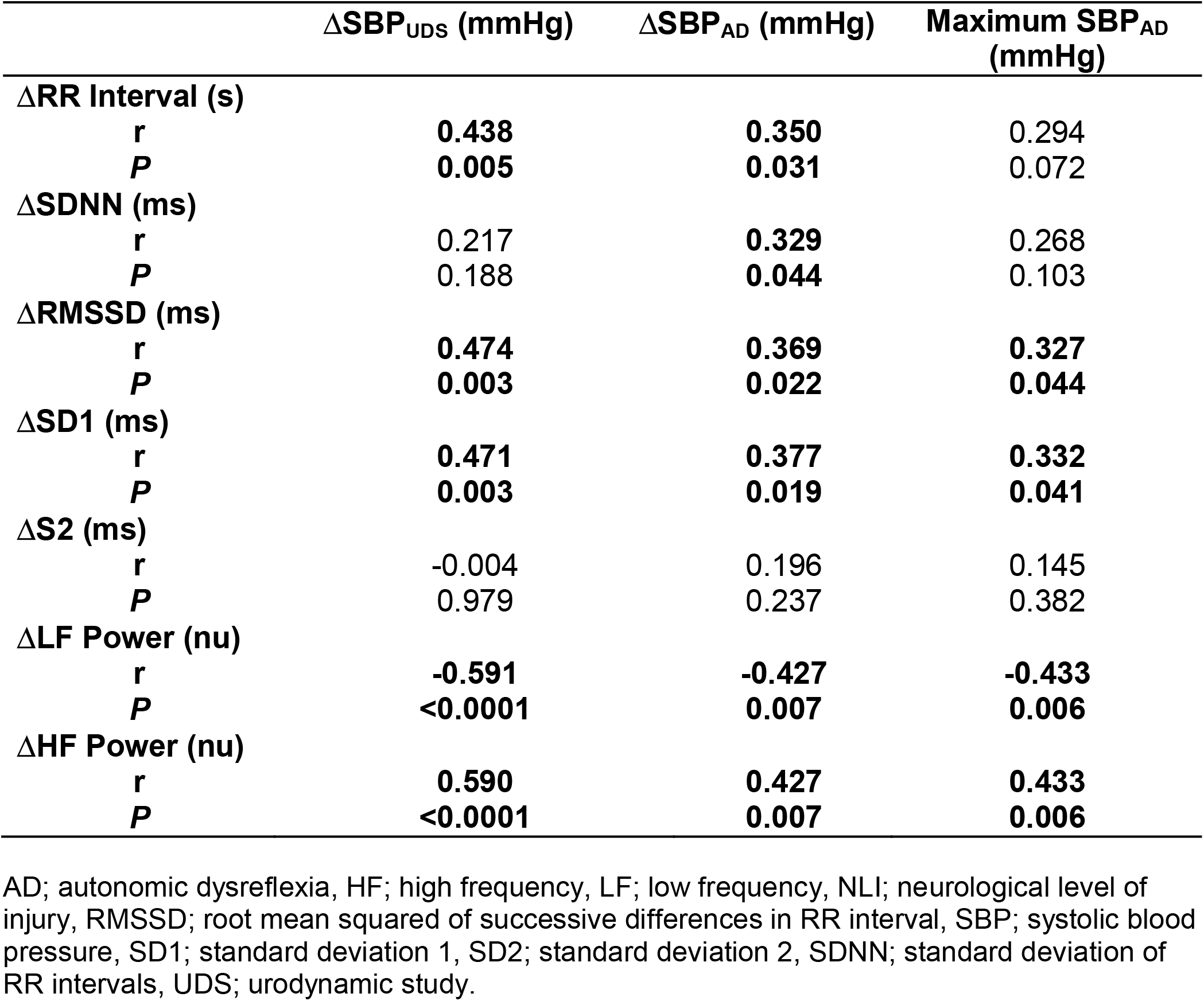
Correlations between time and frequency domain responses to UDS and the change in SBP during UDS (ΔSBP_UDS_), AD (ΔSBP_AD_), and maximal SBP during AD.

## DISCUSSION

In the present study, we demonstrate that bladder filling during UDS in individuals with chronic SCI at T6 or above is associated with increased BP that results in a parasympathetic response characterized by increased RR-interval and HRV. Furthermore, we present evidence that intradetrusor onabotulinumtoxinA injections significantly reduce the responses to bladder filling during UDS in this cohort. These novel findings are in line with the beneficial effects of intradetrusor onabotulinumtoxinA injections that have been reported in the SCI population.^28^

Regulation of LUT function involves a complex interplay between voluntary motor control and involuntary autonomic control of the central nervous system.^29,30^ In healthy individuals, distention of the urinary bladder results in sympathetic activation and parasympathetic innervation of the detrusor muscles is inhibited.^29,31^ This prevents involuntary bladder emptying and has shown to result in minor increases in HR and BP as bladder filling increases.^32,32^ In line with this, Mehnert *et al*. have shown that in healthy volunteers, HR and LF power both increase throughout bladder filling during UDS while HF power decreases.^33^ However, in SCI, loss of descending motor and autonomic control of the bladder causes NDO that is mediated by spinal reflex pathways.^1,29^ Distention of the urinary bladder sends afferent sympathetic signals that are sustained in spinal reflex pathways during bladder filling which cause vasoconstriction below the level of injury and result in elevated BP and AD.^1,29^ Here, we show that bladder filling during UDS results in a parasympathetic response as seen by increases in RR-interval and well-defined HRV markers of parasympathetic activity such as RMSSD, SD1, and HF power. We hypothesize that this is the result of cardiovagal baroreflex activation secondary to elevated BP during bladder filling. Consistent with this idea, there was a significant association between the change in these markers during bladder filling and the change in SBP at the same timepoint.

Contrary to our findings, two previous studies examined the relationship between UDS and HRV in individuals with SCI and found that HRV did not change in response to bladder filling.^34,35^ Neither study saw any changes in BP during UDS which likely accounts for the lack of an observed effect on HRV. Furthermore, the study by Gomez *et al*. recruited predominantly individuals with thoracic SCI.^34^ We observed notable differences in how cervical vs thoracic SCI participants responded to bladder filling during UDS. Individuals with thoracic SCI appeared to have minimal changes in HRV in response to bladder filling during UDS while cervical SCI participants demonstrated robust increases in RR-interval as well as both time and frequency domain HRV. This is consistent with previous studies showing increased severity of autonomic dysfunction and increased HF power in individuals with cervical SCI compared to thoracic SCI.^16^ These data may suggest that in-tact sympathetic pathways in thoracic SCI are able to effectively oppose reflex vagal activation of the sinoatrial node during UDS.

Injection of onabotulinumtoxinA into the detrusor results in block of the pre-synaptic release of acetylcholine from the parasympathetic innervation to produce partial paralysis of the detrusor muscle.^29^ This in turn results in minimized reflexive sympathetic activation of spinal reflex pathways and lowers SBP responses to bladder filling.^12^ Consequently, lower SBP responses to bladder filling cause diminished activation of the cardiovagal baroreflex.^16^ Consistent with our findings demonstrating that treatment had no effect on HRV at rest, Mehnert *et al*.^36^ have published similar findings showing intradetrusor onabotulinumtoxinA injections had no impact on time or frequency domain parameters in participants with NDO. Despite this, their study did not examine how HRV changed under physical stress, such as UDS. Accordingly in the present study, HRV analysis post-treatment revealed reduced reflex vagal responses to bladder filling as evident by reduced HF power during UDS. We also found a significant association between the change in frequency domain metrics of HRV, RMSSD and SD1 during bladder filling and the severity of AD. These data may provide some preliminary evidence for the use of HRV monitoring during UDS in order to predict AD. However, because this study is a secondary analysis, we acknowledge that the sample size may not be adequately powered to determine the sensitivity and specificity of HRV as a predictor of AD.

This study highlights the potential for intradetrusor onabotulinumtoxinA injections to minimize reflex autonomic responses associated with UDS in this cohort. Our study does however have some limitations. It is well known that respiratory rate can have a significant effect of HRV and was not controlled in the present study. While this may have a subtle influence on variability between participants, the pre-post design of the study compares outcomes within participants who’s breathing patters are likely to remain constant from day-to-day. Furthermore, the robust and significant effects of our intervention suggest that the impacts of UDS on HRV are larger than those imposed by alterations in respiratory rate.

We present two novel findings in the present study which show that UDS increases reflex parasympathetic activation, as assessed by HRV, in response to bladder filling during UDS and that intradetrusor onabotulinumtoxinA injections ameliorate this response in individuals with SCI. Furthermore, we provide evidence that HRV may be a useful monitoring tool for early prediction of AD during UDS.

## Data Availability

The data are available from the corresponding author on reasonable request.

## Author Contributions

T.W.D., M.W., and A.V.K. contributed to conception and experimental design. M.W. and A.V.K. acquired the data. T.W.D. analyzed and interpreted the data. T.W.D. drafted the manuscript. T.W.D., M.W., and A.V.K. contributed to editing the final version of the manuscript. M.W. and A.V.K. supervised the project.

